# Weighing psychosocial factors in relatives for the risk of depressive and anxious psychopathology: A sibling-pair comparison study

**DOI:** 10.1101/2022.11.04.22281939

**Authors:** Eleonore D. van Sprang, Dominique F. Maciejewski, Yuri Milaneschi, Marie-Louise J. Kullberg, Bernet M. Elzinga, Albert M. van Hemert, Catharina A. Hartman, Brenda W. J. H. Penninx

## Abstract

**Purpose:** Siblings of probands with depressive and anxiety disorders are at increased risk for psychopathology, but little is known about how risk factors operate within families to increase psychopathology for siblings. We examined the additional impact of psychosocial risk factors in probands – on top of or in combination with those in siblings – on depressive/anxious psychopathology in siblings.

**Methods:** The sample included 636 participants (*M*_age_= 49.7; 62.4% female) from 256 families, each including a proband with lifetime depressive and/or anxiety disorders and their sibling(s) (*N*=380 proband-sibling pairs). Sixteen psychosocial risk factors were tested. In siblings, depressive and anxiety disorders were determined with standardized psychiatric interviews; symptom severity was measured using self-report questionnaires. Analyses were performed with mixed-effects models accounting for familial structure.

**Results:** In siblings, various psychosocial risk factors (female gender, low income, childhood trauma, poor parental bonding, being single, smoking, hazardous alcohol use) were associated with higher symptomatology and likelihood of disorder. The presence of the same risk factor in probands was independently associated (low income, being single) with higher symptomatology in siblings or moderated (low education, childhood trauma, hazardous alcohol use) – by reducing its strength – the association between the risk factor and symptomatology in siblings. There was no additional impact of risk factors in probands on likelihood of disorder in siblings.

**Conclusion:** Our findings demonstrate the importance of weighing psychosocial risk factors within a family context, as it may provide relevant information on the risk of affective psychopathology for individuals.

## Introduction

One of the strongest risk factors for the onset of depressive and anxiety disorders is a family history of these disorders [1, 2]. A two- to three-fold increased risk of the disorders is found for siblings of probands with depressive and anxiety disorders as compared to individuals without affected relatives [3–5]. However, despite their increased risk for psychopathology, siblings from the same at-risk family can differ substantially from one another in psychological functioning [6, 7]. Consistent with this, we showed in a previous study that proband-sibling resemblance in several psychopathology-related features (i.e. symptoms, social/cognitive vulnerabilities, personality traits) was only mild to moderate [8]. Although a large body of evidence exists for the association between several psychosocial risk factors (e.g. female gender, socioeconomic deprivation, social isolation, poor parental bonding, adverse events, smoking, alcohol (ab)use, physical inactivity) and high risk for depressive and anxiety disorders [9–21], little is known about how these risk factors operate within families to increase psychopathology for at-risk siblings. Siblings’ increased risk for psychopathology may depend on the presence of such risk factors in their affected proband, either by also being present in the proband (e.g. an additional ‘vicarious’ effect) [22] or by being absent in the proband while being present in the sibling (e.g. the feeling of ‘being the black sheep’ in the family) [23–25]. This may even extend to sociodemographic risk factors such as higher age and female gender: for instance, rumination seems to be ‘contagious’ especially among older and same-sex female sibling pairs potentially due to the stronger emotional bonds and social learning/sharing [26–29]. Co-rumination, in turn, has been found to be associated with affective psychopathology [30, 31]. Identifying how psychosocial risk factors of poor mental health in siblings of affected probands operate within families may help identifying potential mechanisms explaining why some siblings develop a depressive and anxiety disorder, whereas others do not.

While a large number of studies have investigated the familial aggregation of depressive and anxious psychopathology, only a few have examined the impact of not only considering psychosocial risk factors within at-risk siblings, but also within their affected proband, on increasing psychopathology in at-risk siblings. Findings were mixed as to whether the individual risk for psychopathology is increased if risk factors in a relative are *also present* or if risk factors in a relative are *absent*. A larger neighborhood socio-economic deprivation [32], higher childhood emotional maltreatment [33], and poorer parental bonding [34–37] in an individual as compared to their sibling(s) was found to be associated with more severe depressive symptoms of that individual. Results were mixed for age and gender, with some studies finding associations of similarity (vs. dissimilarity) in female gender and age with similarity in depressive/anxious psychopathology [4, 38], while other studies found no added impact of taking into account the gender/age of an individual’s sibling for their risk for psychopathology [5, 39, 40]. Moreover, available studies have mainly been limited to investigate the degree but not the direction of proband-sibling (dis)similarity of risk factors [4, 5, 38, 40, 41] and mainly focused on sociodemographic and early life adversity risk factors, but not on a wider variety of psychosocial risk factors (e.g. also including recent life adversity and lifestyle-related factors).

The present study examined how a broad range of established psychosocial risk factors for depression/anxiety operate within families to explain interindividual differences in psychopathology between siblings of probands with depressive/anxiety disorders. The main aim was to disentangle and quantify the effect of the presence of a risk factor in the proband, by testing whether this (i) had a unique contribution for psychopathology in the sibling, over-and-above the presence of this risk factor in the sibling, and/or (ii) modified the association between this risk factor and psychopathology in the sibling.

## Methods

### Study sample

Participants were from the Netherlands Study of Depression and Anxiety (NESDA), an ongoing longitudinal cohort study (2004-present) investigating the long-term course and consequences of depressive and anxiety disorders. A detailed description of the NESDA study design and sampling procedure has been reported elsewhere [42]. During the 9-year follow-up (2014-2017), full-biological siblings of NESDA participants with a lifetime depressive and/or anxiety disorder were additionally recruited for the NESDA family study (see Van Sprang et al. [8] for inclusion criteria and sampling procedure). The study sample included 636 participants from 256 unique families: 256 lifetime affected probands and their 380 siblings (*N* = 380 proband-sibling pairs). The present study used data for probands assessed at the 9-year follow-up of NESDA, at the time of recruitment and assessment of siblings. The study protocol was approved by the ethical committee of participating universities, and all respondents provided written informed consent.

### Measurements

#### Outcome measures in sibling

The presence of lifetime DSM-IV-TR [43] diagnoses of depressive (i.e. major depressive disorder and dysthymia) and anxiety disorders (i.e. generalized anxiety disorder, panic disorder with or without agoraphobia, social phobia, and agoraphobia only) was determined using the Composite Interview Diagnostic Instrument (CIDI, lifetime version 2.1) [44].

Past week severity and number of symptoms was measured with the Inventory of Depressive Symptomatology-Self Report (IDS) [45] for depression and with the Beck Anxiety Inventory (BAI) [46] for anxiety. As the IDS and BAI showed a large overlap (multilevel^1^ correlation *r* = 0.71, 95% CI 0.67– 0.75, *t*(623) = 25.28, *p* < 0.001), IDS and BAI scores were standardized and averaged into an overall IDS/BAI score to reflect number and severity of current depressive and/or anxiety symptoms.

#### Psychosocial risk factors in probands and siblings

##### Sociodemographics

Sociodemographic risk factors included higher age (in years; i.e. longer exposure time-frame) [47, 48], female gender, low education (i.e. reversed years of education), and low income defined as gross annual income ≤ €33,600 (i.e. income below average in 2014-2017 in the Netherlands) [49].

##### Life adversity and lifestyle

Early life adversity included childhood trauma and poor parental bonding. The Childhood Trauma Questionnaire-Short Form^2^ (CTQ) [50] was used to assess childhood trauma before the age of 16 (subscales: sexual, physical, and emotional abuse, and physical and emotional neglect). The perception of the relationship between participants and their mother (i.e. maternal bonding) and father (i.e. paternal bonding) before the age of 16 was assessed using the shortened 16-item version of the Parental Bonding Instrument (PBI) [51].

Recent life adversity and lifestyle-related risk factors included current unemployment, living alone, being single (i.e. not married or in a steady relationship), small social network, past-year negative life events, smoking status (yes/no), hazardous alcohol use, and physical inactivity. Participants were considered to have a small social network if the total number of relatives, friends, and close acquaintances with whom they have regular and important contact was ≤5. The List of Threatening Experiences (LTE) was used to assess the total number of past-year exposures to two types of negative life events: (i) independent events, which are unlikely to be influenced by the person (e.g. death of a loved one) and (ii) dependent events, which are likely, but do not have to be, influenced by a person (e.g. job loss) [52, 53]. Following the WHO guidelines [54] for the Alcohol Use Disorders Identification Test (AUDIT) [55], hazardous alcohol use was defined as having an AUDIT sum-score ≥8 for participants aged <65 years and, given that the effects of alcohol vary with average body weight and differences in metabolism, as having an AUDIT sum-score ≥7 for participants aged ≥65 years. Physical activity was measured using the Metabolic Equivalent of Task (MET) score, which was derived from the International Physical Activity Questionnaire (IPAQ) [56], and represented the total number of MET-minutes per week of walking, moderate, and vigorous activities divided by 1000. In the analyses, reversed MET-scores were used reflecting risk associated with physical inactivity.

### Statistical analyses

The associations between outcomes in sibling and explanatory variables in siblings and probands were estimated with linear (current symptom severity) and logistic (presence of lifetime psychiatric diagnosis) mixed-effects regressions. All models included a random intercept of ‘Family-ID’ to account for within-family clustering (34.4% of families included more than one proband-sibling pair) [8]. Models with symptom severity in sibling as the outcome were additionally adjusted for symptom severity in the proband^3^.

Analyses were divided in three main steps, separately for each of the 16 risk factors. In Step 1, main effects of risk factors measured in siblings were included as explanatory variables. In Step 2, main effects of the same risk factors measured in probands were added to examine whether there was a unique contribution of this risk in the proband for psychopathology in their sibling(s), over-and-above individual-level sibling risk factors. In Step 3, sibling × proband risk factor interaction terms were added to corresponding Step 2 models to evaluate whether the association between a risk factor and psychopathology in the sibling was moderated by the presence/degree of this risk in the proband. In logistic models the coefficient of the interaction term estimates departure from multiplicativity, rather than departure from additivity as is the case in linear models. Since interaction on the additive scale may reflect biological/psychological interaction better than interaction on the multiplicative scale [57], we additionally tested departure from additivity using the relative excess risk due to interaction (RERI) measure as proposed by Knol et al. [58] for logistic models (information on the calculation of the RERI measure can be found in the supplementary methods). To facilitate the evaluation of the clinical relevance of also taking into account risk factors in probands, percentages of additional explained variance (Δ*R*^*2*^) were reported for risk factors showing a significant proband main effect in Step 2 or a significant sibling × proband interaction in Step 3, as compared to Step 1 (in which only individual-level sibling risk factors were included). In line with recommendations by Nakagawa et al. [59] for *R*^*2*^ in mixed-effects models, both marginal (i.e. additional variance explained by fixed effects) and conditional Δ*R*^*2*^ (i.e. additional variance explained by both fixed and random effects) were reported.

All analyses were performed in R version 3.6.1 [60]. Statistical tests were two-sided and considered to be statistically significant at *p* < 0.05. False discovery rate (FDR) [61] *q*-values^4^ were additionally reported taking into account multiple testing for the total number of tests performed within each analytical step. Proband-sibling pairs with missing data on a variable were deleted listwise from the analyses including that variable.

### Deviations of pre-registration

This paper was pre-registered on the Open Science Framework; here, the R code for the analyses and a detailed description of the deviations from the pre-registered plan can be found as well (https://osf.io/kzq3p/?view_only=a65ae8fac4154d65857773212ede73e5). Briefly, we had initially planned on using proband-sibling difference scores for explanatory variables and outcomes. However, during analyses we realized several problems with this approach with regard to interpretation (e.g. for continuous data, difference scores around zero could mean both high and both low risk for the sibling and their proband, which would likely have different implications for risk for psychopathology). With our new approach, we were able to assess the impact of individual-level sibling and proband risk factors and whether their combination was related to sibling psychopathology.

## Results

The mean age of the sample (*N* = 636) was 49.7 years (*SD* = 13.2, range 20–78), mean years of education was 13.3, and 62.4% were female. Sample characteristics for probands and siblings separately can be found in Table 1. Of the 380 siblings, 191 (50.3%) had a lifetime depressive and/or anxiety disorder diagnosis. Missing data on study variables was small (Supplementary Table 1). Pairwise multilevel correlations between psychosocial risk factor variables can be found in Supplementary Table 2.

**Table 1.** Sample characteristics of probands and siblings

|  | Probands<br>*N* = 256 | Siblings<br>*N* = 380 |
| --- | --- | --- |
| **Sociodemographics** |  |  |
| Age (years), *M* (*SD*) | 48.52 (13.10) | 50.46 (13.25) |
| Female gender, % | 73.4 | 55.0 |
| Education (years), *M* (*SD*) | 13.42 (2.99) | 13.17 (3.22) |
| Low income, % | 52.9 | 47.5 |
| **Life adversity and lifestyle** |  |  |
| **Early life** |  |  |
| Childhood trauma, *M* (*SD*) | 38.59 (10.64) | 37.18 (9.54) |
| Poor parental bonding – Maternal, *M* (*SD*) | 31.70 (8.88) | 30.63 (8.26) |
| Poor parental bonding – Paternal, *M* (*SD*) | 31.90 (8.57) | 31.20 (8.26) |
| **Recent life** |  |  |
| Unemployment, % | 36.7 | 29.6 |
| Living alone, % | 28.9 | 20.8 |
| Being single, % | 25.4 | 21.3 |
| Small social network, % | 38.6 | 24.5 |
| Negative life events – Independent, *M* (*SD*) | 0.30 (0.60) | 0.45 (0.67) |
| Negative life events – Dependent, *M* (*SD*) | 0.21 (0.54) | 0.27 (0.59) |
| Smoking status, % | 20.3 | 24.3 |
| Hazardous alcohol use, % | 30.8 | 28.5 |
| Physical activity, *M* (*SD*) | 3.58 (2.92) | 4.15 (3.48) |
| **Mental health** |  |  |
| Current depressive symptom severity, *M* (*SD*) | 16.45 (10.49) | 13.15 (9.72) |
| Current anxiety symptom severity, *M* (*SD*) | 9.22 (8.11) | 5.73 (6.09) |
| Current depressive and/or anxiety disorder diagnosis, % | 37.5 | 26.8 |
| Lifetime depressive and/or anxiety disorder diagnosis, % | 100.0 | 50.3 |

*Note.* Sample sizes vary slightly due to marginally missing data on psychosocial risk factors (Supplementary Table 1). *M* = mean; *SD* = standard deviation.

Associations of explanatory variables in siblings and probands and outcomes in siblings are reported in Table 2 (current symptom severity; linear mixed models) and Table 3 (lifetime psychiatric diagnosis; logistic mixed models). Analyses with individual-level sibling risk factors only (Step 1; Table 2) showed that more severe symptoms in the sibling were associated with female gender (*γ* = 0.33, *SE* = 0.08, *p* < 0.001), low income (*γ* = 0.39, *SE* = 0.08, *p* < 0.001), unemployment (*γ* = 0.32, *SE* = 0.09, *p* < 0.001), being single (*γ* = 0.24, *SE* = 0.10, *p* = 0.020), smoking (*γ* = 0.21, *SE* = 0.10, *p* = 0.030), hazardous alcohol use (*γ* = 0.26, *SE* = 0.09, *p* = 0.006), higher levels of childhood trauma (*γ* = 0.34, *SE* = 0.04, *p* < 0.001), and poorer maternal (*γ* = 0.25, *SE* = 0.04, *p* < 0.001) and paternal bonding (*γ* = 0.27, *SE* = 0.04, *p* < 0.001). These risk factors were also significantly associated with an increased likelihood of lifetime psychiatric diagnosis in the sibling (all *p* < 0.05; Step 1; Table 3), except for unemployment (OR = 1.64, *SE* = 0.42, *p* = 0.051).

**Table 2.**
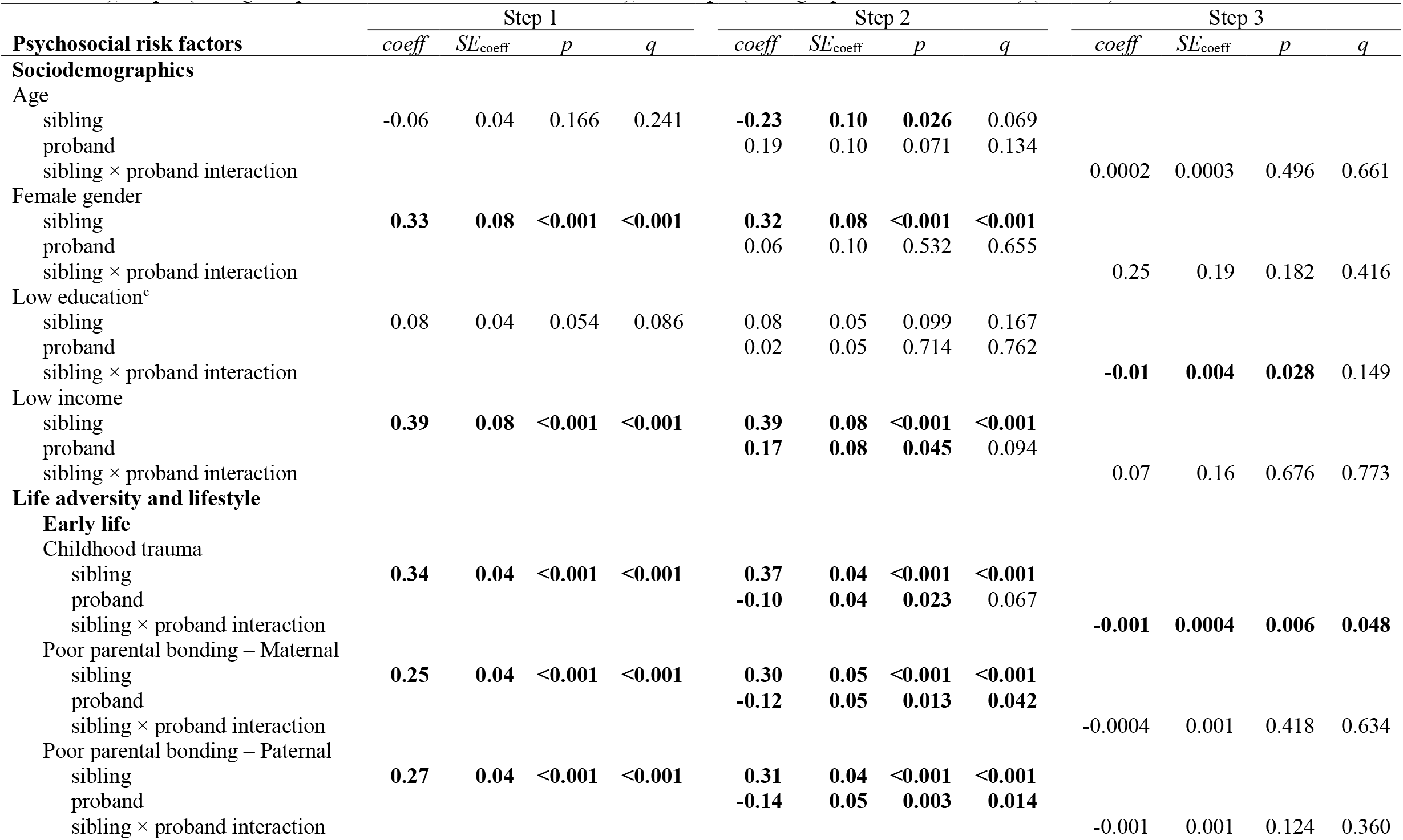
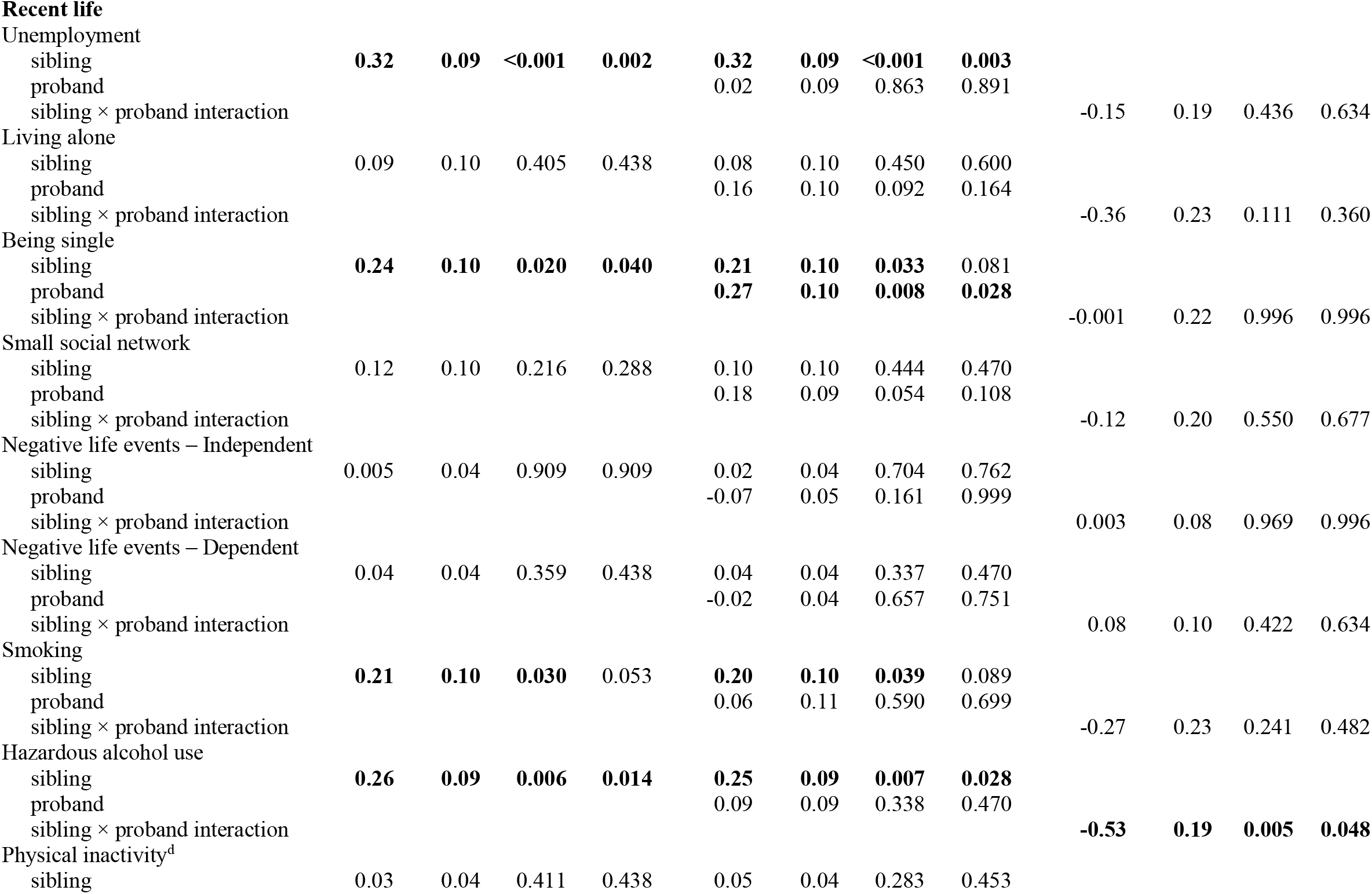
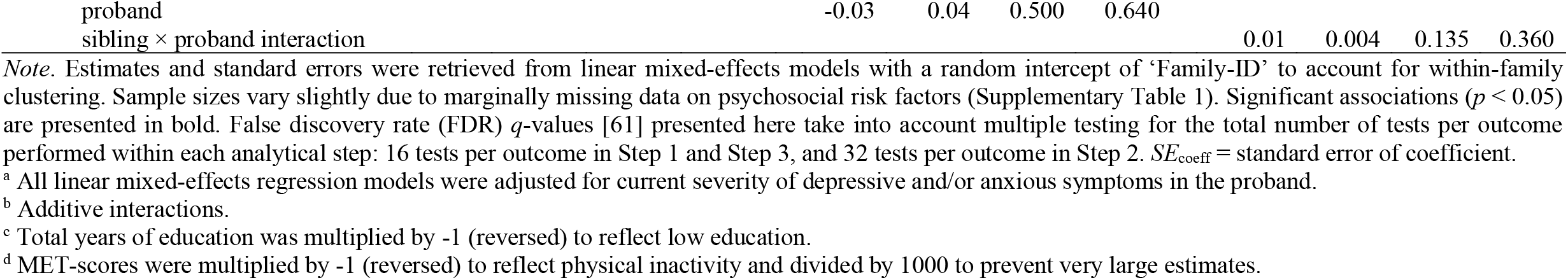
Adjusted^a^ associations of psychosocial risk factors with current depressive and/or anxious symptoms in the sibling: Step 1 (sibling individual-level associations), Step 2 (sibling and proband individual-level associations), and Step 3 (sibling × proband interactions^b^) (*N* = 380)

**Table 3.**
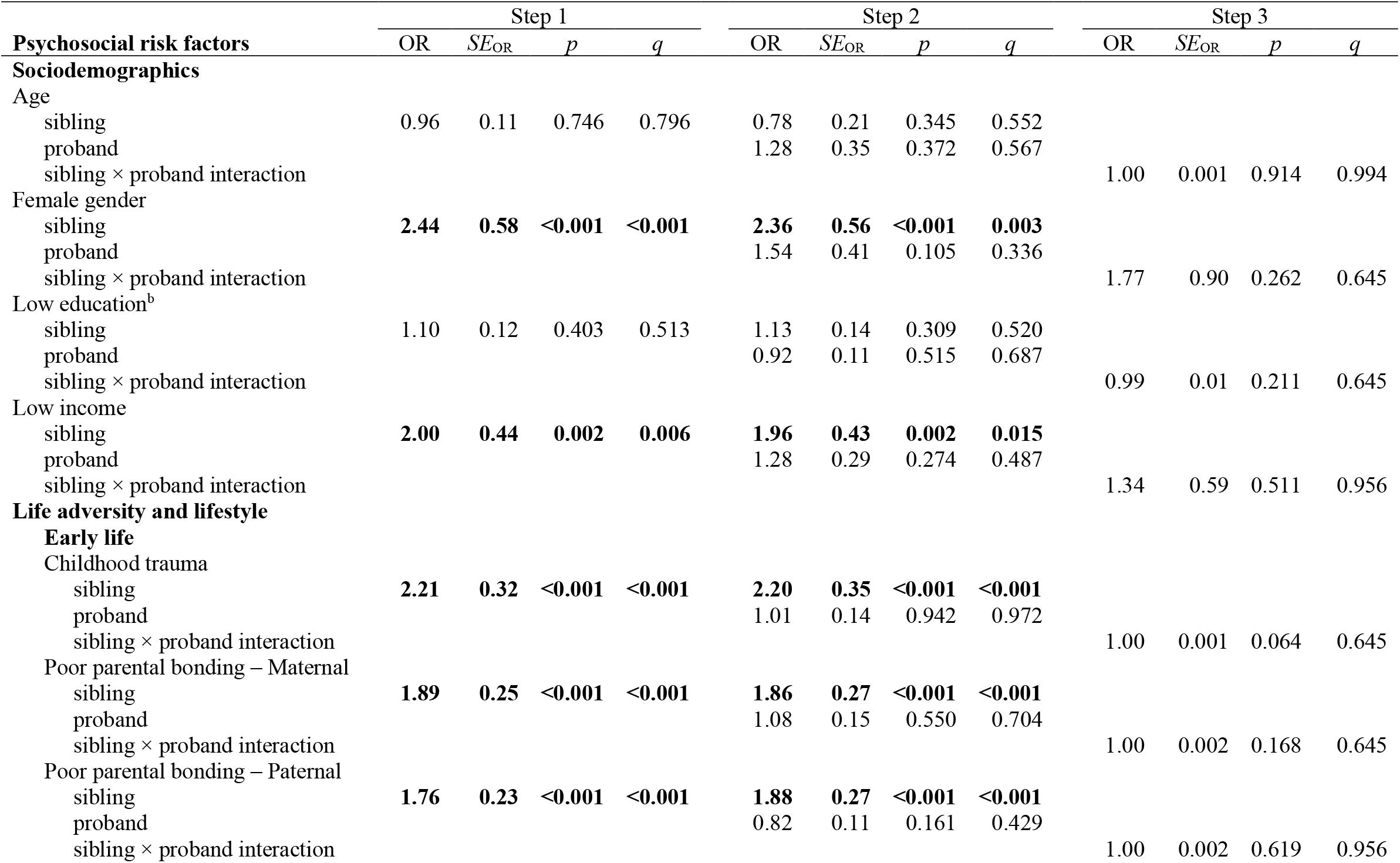
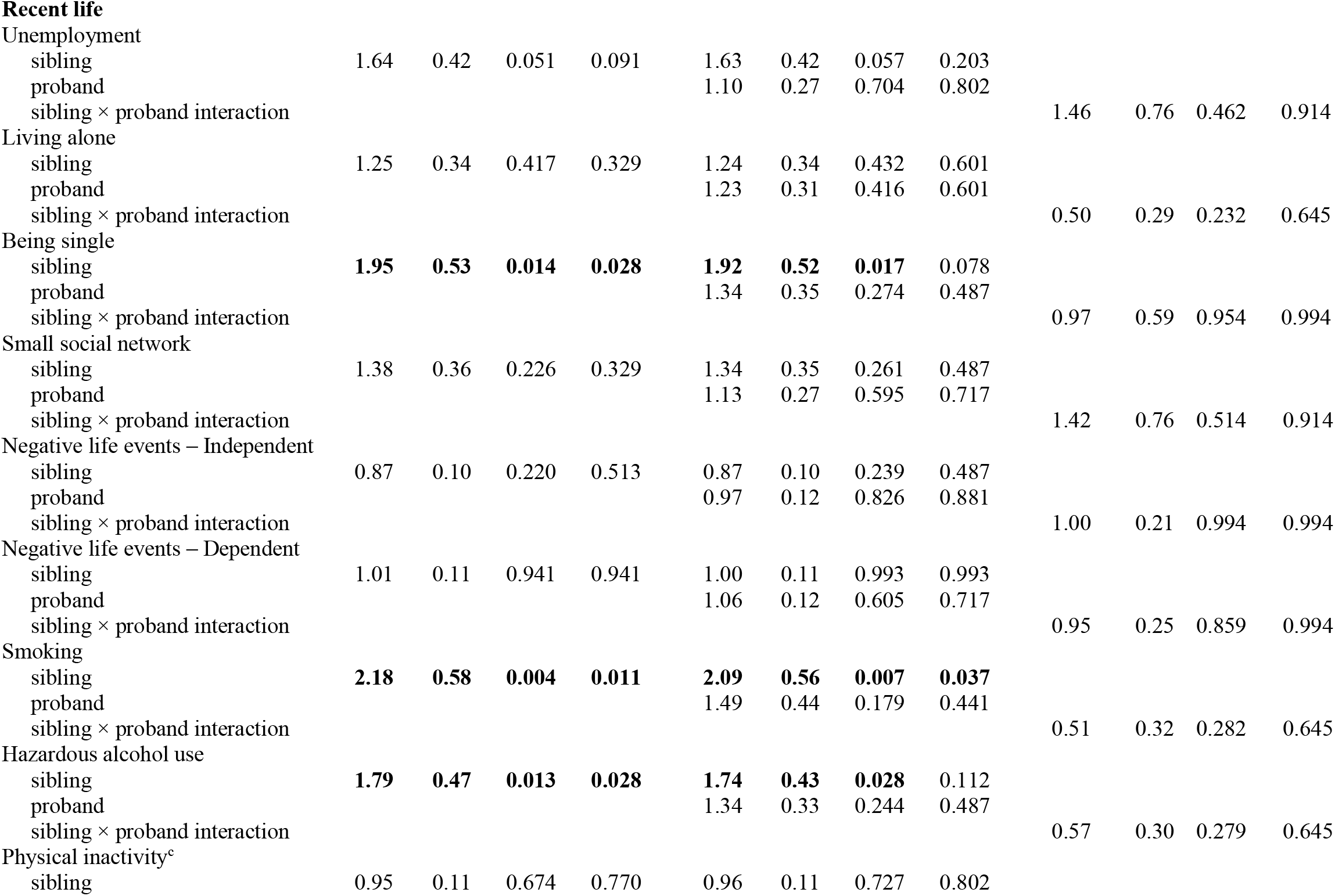
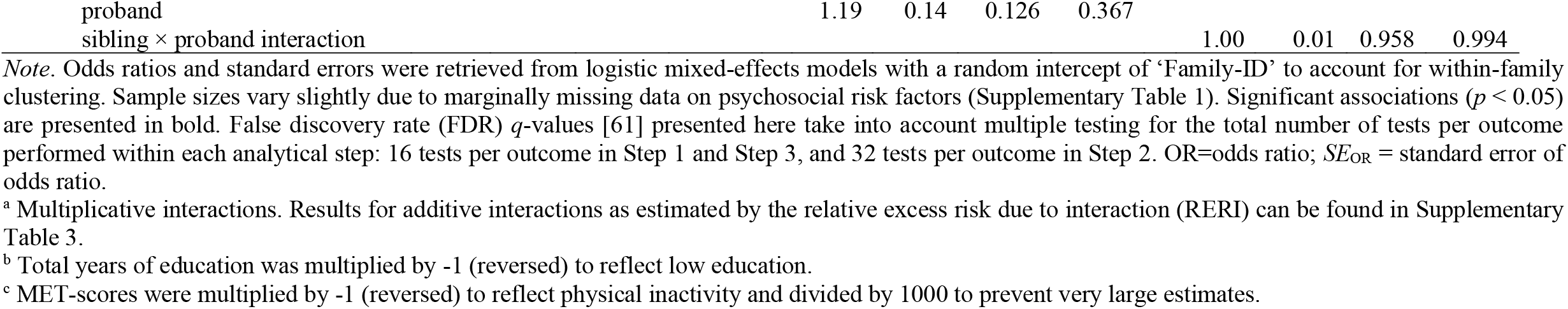
Associations of psychosocial risk factors with lifetime depressive and/or anxious psychopathology in the sibling: Step 1 (sibling individual-level associations), Step 2 (sibling and proband individual-level associations), and Step 3 (sibling × proband interactions^a^) (*N* = 380)

In Step 2, we added the same risk factors assessed in probands as additional explanatory variables. For the outcome of current symptomatology in sibling, main effects of several proband individual-level risk factors were found (Step 2; Table 2): on top of the presence/degree of the risk factor in the sibling, proband low income (*γ* = 0.17, *SE* = 0.08, *p* = 0.045, Δ*R*^*2*^_*marginal*_ = 0.4%, Δ*R*^*2*^_*conditional*_ = 0.7%), being single (*γ* = 0.27, *SE* = 0.10, *p* = 0.008, Δ*R*^*2*^_*marginal*_ = 1.8%, Δ*R*^*2*^_*conditional*_ = 3.4%), and lower levels of childhood trauma (*γ* = -0.10, *SE* = 0.04, *p* = 0.023, Δ*R*^*2*^_*marginal*_ = 0.2%, Δ*R*^*2*^_*conditional*_ = 2.7%), and more optimal maternal (*γ* = -0.12, *SE* = 0.05, *p* = 0.013, Δ*R* ^*2*^_*marginal*_ = 1.5%, Δ*R*_*conditional*_^*2*^ = 1.5%) and paternal bonding (*γ* = -0.14, *SE* = 0.05, *p* = 0.003, Δ*R*^*2*^_*marginal*_ = 2.1%, Δ*R*^*2*^_*conditional*_ = 1.2%) were associated with more severe symptoms in the sibling. No significant main effects of proband risk factors on the outcome of lifetime psychiatric diagnosis in sibling were found (all *p* > 0.05; Step 2; Table 3).

The additional effect of sibling × proband interactions in risk factors was tested in Step 3. For the additive interaction effect on current symptomatology in the sibling (Step 3; Table 2), significant sibling × proband interactions were found for low education (*γ* = -0.01, *SE* = 0.004, *p*=.028, Δ*R*^*2*^_*marginal*_= 1.3%, Δ*R*^*2*^_*conditional*_ = 1.1%), childhood trauma (*γ* = -0.001, *SE* = 0.0004, *p* = 0.006, Δ*R*^*2*^_*marginal*_ = 1.8%, Δ*R*^*2*^_*conditional*_ = 0.7%), and hazardous alcohol use (*γ* = -0.53, *SE* = 0.19, *p* = 0.005, Δ*R*^*2*^_*marginal*_ = 2.2%, Δ*R*^*2*^_*conditional*_ = 2.4%). Fig. 1 shows the association between a risk factor and symptoms in the sibling for different values of that risk factor in the proband for low education (left panel), childhood trauma (middle panel), and hazardous alcohol use (right panel). Consistently, when the risk factor was also present in the proband (lower years of education, higher levels of childhood trauma, and hazardous alcohol use), the strength of the association between the same risk factor and symptoms in their sibling was reduced. No significant (multiplicative) sibling × proband interactions were found for the outcome of lifetime psychiatric diagnosis in sibling (all *p* > 0.05; Step 3; Table 3), nor when the coefficient of the interaction term estimated departure from additivity (Supplementary Table 3).

**Fig. 1.**
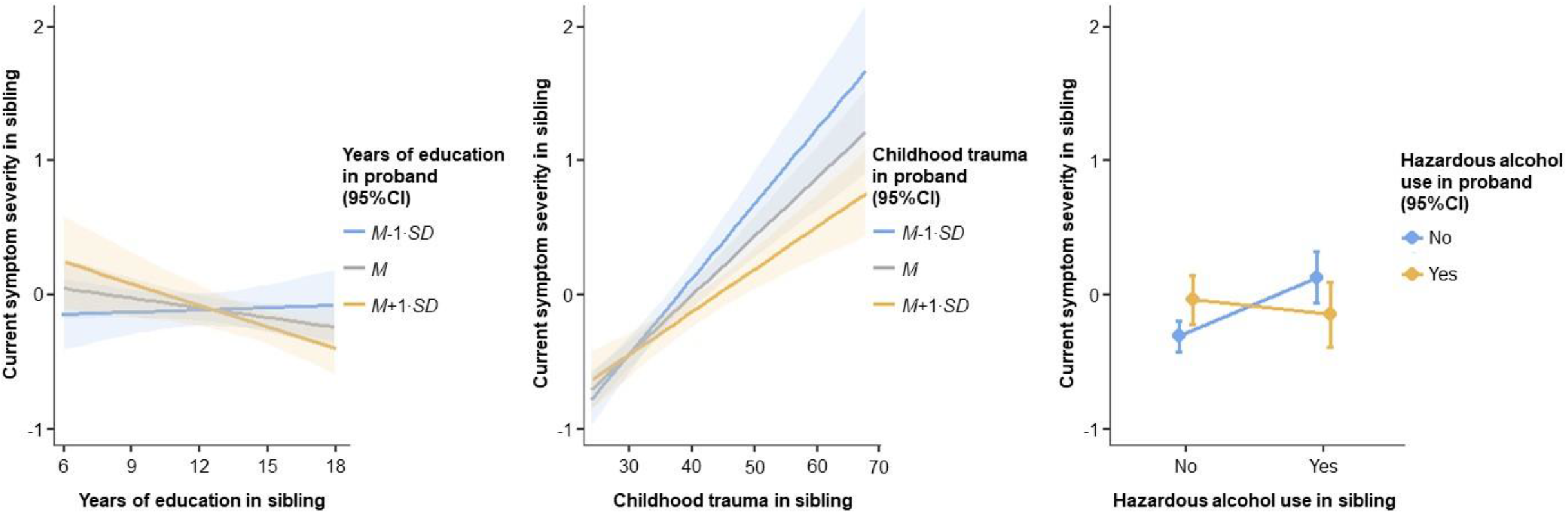
Fixed effects (with 95% confidence intervals) of sibling × proband interaction effects of years of education (left panel, childhood trauma (middle panel), and hazardous alcohol use (right panel) on current depressive and/or anxiety symptom severity in the sibling, while controlling for current symptomatology in the proband. Current depressive and/or anxiety symptom severity was measured as standardized and averaged overall IDS/BAI score. Estimates of simple effects (γ) were retrieved from linear mixed-effects models with a random intercept of ‘Family-ID’ to account for within-family clustering and indicate the (presence/absence and direction of) association between a risk factor and current symptomatology in the sibling for different values of that risk factor in the proband. Low years of education (left panel) was associated with more severe symptoms in the sibling when the proband had high years of education (*M* + 1·*SD*: *γ* = -0.05, *SE* = 0.02, *p* = 0.007); no associations were found when the proband had average (*M*: *γ* = -0.02, *SE* = 0.01, *p* = 0.104) or low years of education (*M* - 1·*SD*: *γ* = 0.01, *SE* = 0.02, *p* = 0.781). Higher childhood trauma (middle panel) was associated with more severe symptoms in the sibling and the strength of this association increases for decreasing trauma levels in the proband (*M* + 1·*SD*: *γ* = 0.03, *SE* = 0.01, *p* < 0.001; *M*: *γ* = 0.04, *SE* = 0.005, *p* < 0.001; *M* - 1·*SD*: *γ* = 0.05, *SE* = 0.01, *p* < 0.001). Hazardous alcohol use (right panel) was associated with more severe symptoms in the sibling when the proband was not a user (No: *γ* = 0.44, *SE* = 0.11, *p* < 0.001); no association was found when the proband was a user (Yes: *γ* = -0.11, *SE* = 0.15, *p* = 0.473). *M* = mean; *SD* = standard deviation; *SE* = standard error.

## Discussion

In siblings of probands with lifetime depressive and anxiety disorders, we confirmed the association of a wide range of established psychosocial risk factors with psychopathology symptoms and disorders. However, the major finding of our study is that the presence of the same risk factors in affected probands, explained additional interindividual differences in psychopathology in at-risk siblings. For instance, siblings having low income and who were single had higher symptoms; intriguingly, the presence of the same risk factor in the proband was additionally associated with higher symptoms in their sibling, independently of the sibling’s individual risk factor. Furthermore, for other risk factors (low education, childhood trauma, hazardous alcohol use), the presence in the proband moderated the association between the risk factor and symptoms in the sibling: when the risk factor was also present in the proband, the strength of the association between the risk factor and symptoms in the sibling was reduced. Thus, when similar levels of a risk factor were shared between probands and siblings, the impact of the risk factor on siblings’ symptoms was buffered. Of note, we only confirmed these additional family effects of risk factors for the continuous outcome of psychopathology symptoms but not for binary clinical diagnoses, possibly due to the reduced statistical power or loss of information when using a dichotomous classification.

Besides showing an additive impact of probands’ low income, which is in line with findings from a previous community-based twin study [32], we extend the current literature by showing that when a proband was single, on top of the presence of the risk in their sibling, this was associated with more severe symptomatology in the sibling. This additive impact of also having low income and being single present in the proband may arise from additional familial clustering between these risk factors and psychopathology (i.e. a more genetic form of the disorders that tends to be co-inherited with these risk factors). In line with this reasoning, twin and genome-wide association studies have shown substantial genetic correlations of income (-0.30 to -0.44) [62, 63] and avoidant/anxious romantic attachment, which might increase the probability of being single, with depressive and anxiety disorders (0.48 to 0.58) [64]. The additive impact of the presence of a risk factor in the proband may also be a reflection of the degree of (and/or other additional) problems within their family. That is, both proband and sibling having low income and being single may indicate a more substantial degree of a family’s socioeconomic deprivation and romantic relationship problems, which likely results from or leads to more severe affective problems [9, 65].

We also found several risk factors for which the additional presence or higher levels in the proband were associated with less severe symptomatology in their sibling(s). These factors included poor parental bonding and childhood trauma, which were also identified in previous studies [33–37, 41], and low education and hazardous alcohol use. Independently of parental bonding levels in the sibling, poorer parental bonding in the proband was associated with less severe symptomatology in the sibling. For childhood trauma, low education, and hazardous alcohol use, the individual-level risk in the proband moderated the impact of the risk factor on symptomatology in their sibling(s): When these factors were similarly present in both proband and sibling, the impact of the factor on sibling symptomatology appeared to be buffered. Conversely, a sibling’s symptoms were higher when these risk factors were not shared with or were of lower level in their proband. This may reflect a ‘black sheep effect’, in which the feeling of having been worse off than your sibling may arise from (perceived) differential parenting (for early life adversity) [66, 67], differences in innate abilities and/or unequal parental resource investment (for education) [68], and sibling deidentification in the proband (i.e. actively seeking to differentiate themself from their sibling; for alcohol use) [69]. Siblings may use each other as a reference point, which in the case of upward social comparisons (i.e. comparisons to a perceived ‘superior’ other) may lead to experiences of unfairness and inequity [24]. Such upward social comparisons have been shown to have the most detrimental effects on depressive and anxious psychopathology by feeding into dysfunctional beliefs about the self [25]. Of note, among risk factors for which we found significant additional impact of considering proband levels, evidence was less strong for low income/education given that these associations were not significant after correcting for multiple testing.

We did not find evidence for an added effect of taking into account proband risk over-and-above or in combination with a sibling’s individual risk for any of the other psychosocial risk factors that we tested, which is in line with previous findings (age, gender) [5, 39, 40]. For instance, recent negative life events, for which we reported moderate proband-sibling resemblance in a previous study [8], were not associated with sibling psychopathology. One possible reason for that is that we measured recent life events in a sample of relatively older aged adults, in which factors beyond the family environment (such as individual, rather than familial, recent negative life events) may have a larger impact. As such cross-sibling effects of recent negative life events on affective psychopathology may be less likely. This is in line with evidence from behavioral-genetic research [6, 7] suggesting an increased role across the lifespan for individual environments and unique risk and protective factors in shaping behavioral, psychological, and personality features.

Overall, results were highly similar between the two outcomes with regard to the direction of associated risk factors, but lifetime diagnosis (dichotomous) showed fewer associated factors within siblings and no associated proband main effects or sibling × proband interaction effects for any of the risk factors, as compared to current symptomatology (continuous). This suggests that the continuous outcome may have provided deeper resolution and/or higher statistical power. This is particularly the case with regard to recent life risk factors and the dichotomous outcome, since we investigated associations between a risk factor that occurred in current/recent life (e.g. unemployment) with a disorder that may have occurred years before the time of assessment, whereas the continuous outcome referred to current symptoms. This may have reduced power to detect proband main effects and/or sibling × proband interaction effects of recent life risk factors and the dichotomous outcome by diluting effects in both siblings and probands.

Strengths of the present study include the sibling structure of the data, which has the advantage that sibling relationships contain a higher shared proportion of (early) environmental factors as compared to parent-offspring relationships; the relatively older age of the sample, which allows for the examination of siblings’ more definite clinical profiles and interindividual discrepancies between siblings that emerged across the lifespan; and the wide variety of assessed psychosocial risk factors. However, the present study is not without limitations. First, as this study only used cross-sectional data, no conclusion can be drawn with regard to the ordering of effects. In particular with regard to some of the lifestyle risk factors, such as smoking or hazardous alcohol use, the association with depressive and anxious psychopathology may be bidirectional [19, 20, 70]. Second, although this study used a relatively large clinically relevant sibling sample of 380 proband-sibling pairs, we may have had insufficient power to detect the examined sibling × proband risk factor interaction effects: based on the assumption that the interaction effect is half size of the main effects, 16 times the sample size is required to estimate an interaction than to estimate a main effect [71]. Third, retrospective self-report measures of life adversity may have been confounded by participants’ differential recall accuracy and current mood. However, we deem the impact low because previous NESDA and other studies showed that these measures had adequate temporal stability and were not critically affected by respondents’ current mood [72–78]. Fourth, this study explored as a ‘bench-mark’ the added impact of individual psychosocial risk factors in probands, over-and-above or in combination with risk factors in siblings. Given the relatively exploratory nature of our study and the fact that several of the studied risk factors were correlated and may therefore explain overlapping portions of the symptom variance, we used separate analytical models for each risk factor. For future research it would definitely be worthwhile to investigate the impact of individual risk factors in probands on top of a broad set of risk factors in siblings. Fifth, the fact that the present study was designed to include a high-risk sample of probands with a lifetime depressive and/or anxiety disorder and their siblings limits the generalizability of the findings to the general population.

To conclude, this study confirmed the association of a broad range of psychosocial risk factors with depressive and anxiety symptoms and disorders in siblings of probands with a lifetime depressive or anxiety disorder. However, importantly, we demonstrated that not only the risk factors within these at-risk individuals are important, but also those within their relatives: the individual-level risk factor in the proband, in itself or in combination with the individual-level risk factor in their sibling, had additional value for siblings’ psychopathology over only considering the individual-level risk factor in the sibling. Even though percentages of additional explained variance were small, previous research [79] has argued that small effects are the norm, rather than the exception, and form an indispensable foundation for cumulative psychological science: small effects may still have substantial direct consequences for individual mental well-being, especially for effects that accumulate over time and at scale such as childhood trauma [80, 81] and low income [82]. Our findings underscore the importance of weighing risk factors within a family context, as whether or not risk factors are shared with other siblings in the family may provide relevant information on the individual risk of depressive/anxious psychopathology. Future studies are needed to identify the exact mechanisms explaining the additive impact of weighing risk factors within all siblings in the family. Moreover, given the recent findings of little specificity in familial transmission in specific classes of psychiatric disorders [83–85], future studies may want to examine whether our findings extend to other risk factors and/or psychiatric conditions.

## Supporting information

Supplemental methods and results

STROBE checklist

## Data Availability

The data that support the findings of this study are available via the website of NESDA (https://www.nesda.nl/pro-index/), which will be provided after handing in a data request. This paper was pre-registered on the Open Science Framework; here, the R code for the analyses can be found as well (https://osf.io/kzq3p/?view_only=a65ae8fac4154d65857773212ede73e5).

## Declarations

### Author contributions

B.W.J.H.P. developed the NESDA study concept and design. Together with B.W.J.H.P., B.M.E., and A.M.v.H. were closely involved in the design of the NESDA family study. E.D.v.S. and M.K. prepared the data for the analyses. E.D.v.S. performed the data analysis and interpretation under supervision of D.F.M., Y.M., and B.W.J.H.P. E.D.v.S. drafted the manuscript, and B.W.J.H.P., D.F.M., Y.M., M.K., C.A.H., B.M.E., and A.M.v.H. provided critical revisions. All authors approved the final version of the paper for submission.

### Ethics approval

The NESDA study protocol was approved by the ethical committee of participating universities. The procedures used in this study adhere to the tenets of the Declaration of Helsinki.

## Footnotes

1 Unlike a normal Pearson correlation coefficient, a multilevel Pearson correlation coefficient takes into account the within-family clustering of the IDS and BAI data (34.4% of families in the present study included more than one proband-sibling pair) [8].

2 For probands, CTQ was administered at the 6-year follow-up.

3 This was done to rule out the possibility that associations were not simply due to familial clustering of depressive/anxious psychopathology. Applying a similar adjustment procedure on models with presence of psychiatric diagnosis in sibling as the outcome was not possible as there was no variance in psychiatric diagnosis in probands (i.e. all probands were lifetime affected).

4 Given the number of statistical comparisons performed within each analytical step (Step 1 and Step 3: 16 comparisons, one for each risk factor, per outcome; Step 2: 32 comparisons, two for each risk factor, per outcome), we deemed it necessary to additionally report FDR-corrected *q*-values. However, uncorrected *p*-values were used as leading in the analyses of this paper: following reasoning by Althouse [86] we believe that adjustment for multiple testing is too strict given the strong prior credibility of the 16 tested risk factors in terms of their association with depressive/anxious psychopathology based on prior research [9–21, 47, 48]. Therefore, uncorrected *p*-values were used as leading, with the sidenote that associations with *p* < 0.05 with but with FDR-corrected *q*≥0.05 should be interpreted with caution.

