## Supplemental methods and results for "Weighing psychosocial factors in relatives for the risk of depressive and anxious psychopathology: A sibling-pair comparison study"

### SUPPLEMENTARY INFORMATION

#### Supplementary methods

##### **Missing data**

Amount of missing data (#, %) for proband and sibling study variables can be found in Supplementary Table 1. All probands (*N* = 256) and siblings (*N* = 380) had complete data on age, gender, years of education, living alone, being single, and lifetime diagnosis of depressive and/or anxiety disorder. All proband-sibling pairs had complete data on age, gender, low education, living alone, and being single. For the remaining study variables, the amount of missing data ranged from 0.3% (*N* = 1) for (for example) low income in the sibling to 8.6% (*N* = 22) for physical inactivity in the proband. Proband-sibling pairs with missing data on a proband or sibling study variable were deleted list wise from the analyses including that variable.

###### **Supplementary Table 1.** Amount of missing data (#, %) proband and sibling study variables

|  | Probands  *N* = 256 | | Siblings  *N* = 380 | |
| --- | --- | --- | --- | --- |
| **Study variables^a^** | *N*_valid_ | *N* (%) missings | *N*_valid_ | *N* (%) missings |
| **Mental health** |  |  |  |  |
| Current severity of depressive symptoms (IDS-SR) | 250 | 6 (2.3) | 376 | 4 (1.1) |
| Current severity of anxiety symptoms (BAI) | 249 | 7 (2.7) | 376 | 4 (1.1) |
| Current severity of depressive and/or anxiety symptoms^b^ | 249 | 7 (2.7) | 376 | 4 (1.1) |
| Current depressive and/or anxiety disorder diagnosis | 256 | 0 (0.0) | 380 | 0 (0.0) |
| Lifetime diagnosis of depressive and/or anxiety disorder | 256 | 0 (0.0) | 380 | 0 (0.0) |
| **Sociodemographics** |  |  |  |  |
| Age | 256 | 0 (0.0) | 380 | 0 (0.0) |
| Female gender | 256 | 0 (0.0) | 380 | 0 (0.0) |
| Low education | 256 | 0 (0.0) | 380 | 0 (0.0) |
| Low income | 255 | 1 (0.4) | 379 | 1 (0.3) |
| **Life adversity and lifestyle** |  |  |  |  |
| **Early life** |  |  |  |  |
| Childhood trauma (CTQ) | 243 | 13 (5.1) | 375 | 5 (1.3) |
| Parental bonding – maternal (PBI) | 249 | 7 (2.7) | 372 | 8 (2.1) |
| Parental bonding – paternal (PBI) | 238 | 18 (7.0) | 363 | 17 (4.5) |
| **Recent life** |  |  |  |  |
| Unemployment | 256 | 0 (0.0) | 379 | 1 (0.3) |
| Living alone | 256 | 0 (0.0) | 380 | 0 (0.0) |
| Being single | 256 | 0 (0.0) | 380 | 0 (0.0) |
| Small social network | 249 | 7 (2.7) | 368 | 12 (3.2) |
| Past-year negative life events – Independent (LTE) | 256 | 0 (0.0) | 379 | 1 (0.3) |
| Past-year negative life events – Dependent (LTE) | 256 | 0 (0.0) | 379 | 1 (0.3) |
| Smoking | 256 | 0 (0.0) | 379 | 1 (0.3) |
| Hazardous alcohol use (AUDIT) | 250 | 6 (2.3) | 376 | 4 (1.1) |
| Physical inactivity (IPAQ) | 234 | 22 (8.6) | 359 | 21 (5.5) |

*Note*. Depressive disorder=major depressive disorder and dysthymia; anxiety disorder = generalized anxiety disorder, panic disorder with or without agoraphobia, social phobia, and agoraphobia; IDS-SR = Inventory of Depressive Symptomatology – Self Report; BAI = Beck Anxiety Inventory; CTQ = Childhood Trauma Questionnaire; LTE = List of Threatening Experiences; AUDIT = Alcohol Use Disorders Identification Test; IPAQ = International Physical Activity Questionnaire.

^a^ Missing items were replaced by the mean of the available items under the condition that the maximum number of missing items was 6 out of 28 for the IDS, 8 out of 21 for the BAI, 5 out of 25 for the CTQ (1 per subscale), 3 out of 16 for the PBI, 1 out of 4 for the LTE – independent events, 1 out of 8 for the LTE – dependent events, and 2 out of 10 for the AUDIT.

^b^ Current depressive and/or anxiety symptom severity was measured as standardized and averaged overall IDS/BAI score.

##### **Calculation of relative excess risk due to interaction (RERI) measure**

Departure from additivity was tested using the relative excess risk due to interaction (RERI) measure [1]. The RERI estimate was deemed to differ significantly from zero if the bootstrapped 95% confidence interval (CI) did not include zero. Bootstrapping was done using the boot package in R with 10,000 bootstrap samples. The outcome of simple, rather than mixed, logistic regression models was used to calculate the RERI and the bootstrapped 95%CI, as bootstrapping of mixed models did not converge (even when using 10 bootstrap samples). As taking into account the family clustering of the data did not significantly improve the model fit, *χ*^2^(1) = 1.46, *p* = 0.226, using simple instead of mixed logistic regression analyses to calculate RERI (and its 95%CIs) is therefore unlikely to have had a substantial impact on model estimates.

#### Supplementary results

##### **Supplementary Table 2.** Pairwise multilevel correlations between psychosocial risk factor variables (N = 636)

|  | 1. | 2. | 3. | 4. | 5. | 6. | 7. | 8. | 9. | 10. | 11. | 12. | 13. | 14. | 15. |
| --- | --- | --- | --- | --- | --- | --- | --- | --- | --- | --- | --- | --- | --- | --- | --- |
| 1. Age (years) |  |  |  |  |  |  |  |  |  |  |  |  |  |  |  |
| 2. Female gender | 0.01 |  |  |  |  |  |  |  |  |  |  |  |  |  |  |
| 3. Education (years) | -0.03 | -0.06 |  |  |  |  |  |  |  |  |  |  |  |  |  |
| 4. Low income | 0.02 | **0.13** | **-0.21** |  |  |  |  |  |  |  |  |  |  |  |  |
| 5. Childhood trauma | -0.02 | **0.10** | -0.08 | **0.13** |  |  |  |  |  |  |  |  |  |  |  |
| 6. Poor parental bonding  – Maternal | 0.02 | **0.15** | -0.07 | **0.12** | **0.53** |  |  |  |  |  |  |  |  |  |  |
| 7. Poor parental bonding  – Paternal | 0.02 | 0.002 | -0.02 | 0.05 | **0.49** | **0.55** |  |  |  |  |  |  |  |  |  |
| 8. Unemployment | **0.23** | **0.09** | **-0.17** | **0.21** | **0.13** | **0.14** | **0.16** |  |  |  |  |  |  |  |  |
| 9. Living alone | 0.06 | -0.02 | 0.02 | **0.43** | 0.06 | 0.05 | 0.01 | **0.13** |  |  |  |  |  |  |  |
| 10. Being single | 0.04 | -0.01 | -0.05 | **0.41** | 0.04 | 0.03 | 0.04 | **0.10** | **0.59** |  |  |  |  |  |  |
| 11. Small social network | **-0.08** | **-0.10** | -0.04 | **0.11** | **0.11** | 0.07 | **0.12** | -0.04 | 0.07 | **0.13** |  |  |  |  |  |
| 12. Negative life events  – Independent | **0.10** | -0.03 | 0.01 | -0.02 | -0.01 | -0.01 | -0.001 | 0.04 | -0.04 | -0.03 | -0.07 |  |  |  |  |
| 13. Negative life events  – Dependent | 0.03 | 0.04 | -0.06 | **0.12** | -0.02 | 0.001 | **-0.10** | **0.13** | **0.13** | **0.08** | -0.06 | **0.08** |  |  |  |
| 14. Smoking status | **-0.11** | **-0.10** | -0.05 | **0.14** | 0.02 | 0.02 | 0.01 | 0.01 | 0.04 | **0.09** | 0.02 | -0.05 | 0.05 |  |  |
| 15. Hazardous alcohol use | -0.04 | **-0.13** | **0.13** | 0.03 | 0.02 | 0.04 | 0.07 | **-0.09** | **0.10** | 0.06 | 0.04 | -0.06 | **0.08** | **0.24** |  |
| 16. Physical inactivity | 0.02 | 0.04 | **0.15** | 0.002 | -0.003 | -0.05 | -0.02 | 0.04 | 0.05 | 0.01 | 0.06 | -0.01 | 0.03 | -0.03 | 0.03 |

*Note*. Multilevel, rather than normal, correlation coefficients were calculated as they take into account the within-family clustering of the data (34.4% of families in the present study included more than one proband-sibling pair) [2]: Pearson correlations between continuous-continuous variable pairs, point-biserial correlations between dichotomous-continuous variable pairs, and tetrachoric correlations between dichotomous-dichotomous variable pairs. Sample sizes vary slightly due to marginally missing data on psychosocial risk factors (Supplementary Table 1). Correlation coefficients significantly different from zero at *p*<.05 are presented in bold.

##### **Supplementary Table 3.** Sibling × proband interactions^a^ of psychosocial risk factors on lifetime depressive and/or anxious diagnosis in the sibling, as reflected by the relative excess risk due to interaction (RERI) (N = 380)

|  | Step 2 | |
| --- | --- | --- |
| **Sibling × proband interactions** | RERI | 95%CI |
| **Sociodemographics** |  |  |
| Age | -0.0002 | -0.003 to 0.001 |
| Female gender | 1.27 | -0.34 to 2.86 |
| Low education | -0.01 | -0.04 to 0.01 |
| Low income | 0.66 | -0.81 to 2.22 |
| **Life adversity and lifestyle** |  |  |
| **Early life** |  |  |
| Childhood trauma | -0.002 | -0.01 to 0.001 |
| Poor parental bonding – Maternal | 0.003 | -0.001 to 0.01 |
| Poor parental bonding – Paternal | -0.002 | -0.01 to 0.002 |
| **Recent life** |  |  |
| Unemployment | 0.58 | -0.82 to 2.49 |
| Living alone | -0.89 | -2.63 to 0.63 |
| Being single | 0.17 | -2.20 to 4.81 |
| Small social network | 0.45 | -0.96 to 2.32 |
| Negative life events – Independent | -0.005 | -0.50 to 0.33 |
| Negative life events – Dependent | -0.05 | -0.84 to 0.81 |
| Smoking | -0.90 | -3.71 to 2.49 |
| Hazardous alcohol use | -0.69 | -2.81 to 1.29 |
| Physical inactivity | 0.0003 | -0.02 to 0.03 |

*Note*. The outcome of simple logistic regression models was used to calculate the RERI and the bootstrapped (using 10,000 bootstrap samples) 95%CI. Sample sizes vary slightly due to marginally missing data on psychosocial risk factors (Supplementary Table 1). RERI = relative excess risk due to interaction; CI = confidence interval.

^a^ Additive interactions.
